# The International Classification of Cognitive Disorders in Epilepsy (IC-CoDE) Portal: An Open Source Resource for Neuropsychological Research in Epilepsy

**DOI:** 10.64898/2025.12.04.25340021

**Authors:** Robyn M. Busch, Tobias Brünger, Kayela Arrotta, Lisa Ferguson, Julie K. Janecek, Sara J. Swanson, Anny Reyes, Carrie R. McDonald, Bruce P. Hermann, Dennis Lal

## Abstract

**Background:** The International Classification of Cognitive Disorders in Epilepsy (IC-CoDE) is a consensus-based, empirically-driven approach to standardize cognitive phenotyping in epilepsy research that has quickly garnered interest within the epilepsy community. However, manually generating IC-CoDE phenotypes in group data is laborious and time-consuming, particularly in datasets containing heterogenous cognitive measures or batteries, limiting the widespread adoption of IC-CoDE phenotypes to further multicenter epilepsy research.

**Methods/Results:** To remove this barrier, we developed the IC-CoDE portal (https://ic-code-portal.ccf.org/), an interactive and user-friendly, web-based platform to support the scientific community in performing IC-CoDE individual level classification through web interface or bulk classification through upload of data files. The portal also allows the user to generate and visualize cohort summary statistics of cognitive phenotypes in their dataset, with the option of filtering the data interactively by specified demographic or clinical variables of interest. We further made the resulting IC-CoDE phenotypes downloadable as a spreadsheet to foster offline analysis of cognitive data by members of the epilepsy research community.

**Significance:** Ascertainment of cognitive profiles is key to understanding the natural heterogeneity in the clinical presentation of the epilepsies and their comorbidities. The IC-CoDE taxonomy can be applied to any comprehensive neuropsychological battery, regardless of the specific test measures and normative data used or the language and culture in which the assessment took place, making it an ideal tool to accelerate international multi-center studies on cognition in epilepsy. We hope that by eliminating some of the barriers associated with cognitive phenotyping, the IC-CoDE portal will springboard large-scale collaborative efforts to further our understanding of the neuropsychology of the epilepsies.

## Introduction

Cognitive impairment is a major comorbidity in epilepsy that can negatively impact day-to-day functioning and quality of life^1^. In fact, individuals with epilepsy report memory and other cognitive difficulties to be among the most concerning aspects of their condition, second only to unexpected seizures and driving restrictions^2^. Understanding the etiology and course of cognitive dysfunction across the lifespan in epilepsy has been a long-standing goal^3^, but these efforts have been hindered by the absence of a standardized diagnostic taxonomy for cognitive disorders in individuals with epilepsy^4^. Cognitive profiling, or phenotyping, is key to understanding the natural heterogeneity in the clinical presentation of a disorder and its comorbidities. This has been clearly demonstrated in the mild cognitive impairment/dementia literature, as well as in a handful of recent epilepsy studies linking cognitive phenotypes to neuroimaging correlates^5–9^.

Recognizing the need for a taxonomy for cognitive disorders in epilepsy to move the field forward, the International League Against Epilepsy (ILAE) Neuropsychology Task Force partnered with the International Neuropsychological Society (INS) in early 2020 to develop the International Classification of Cognitive Disorders in Epilepsy (IC-CoDE), a consensus-based, empirically driven approach to cognitive diagnostics in epilepsy research^4,10^. Importantly, this taxonomy can be applied to any comprehensive neuropsychological battery, regardless of the specific test measures and normative data used or the language and culture in which the assessment took place, making it an ideal tool to further international multi-center studies on cognition in epilepsy. Indeed, this new taxonomy has generated substantial interest in the epilepsy community; it has already been applied to English-speaking and Spanish-speaking patients with focal epilepsies in the U.S. and abroad^10–15^ in patients with epilepsy residing in India^11^ and Germany^16^, to pediatric patients with new onset epilepsy^17^ and drug-resistant epilepsy^18^, to older adults with chronic and late-onset epilepsy^19^, and is the subject of several podcasts and editorials^20,21^.

Unfortunately, while feasible, generating IC-CoDE cognitive phenotypes from a series of standard scores obtained from neuropsychological assessments is currently labor-intensive, cumbersome, and error prone, limiting wide-spread use. To remove this barrier and facilitate large-scale, collaborative international research efforts on cognitive diagnostics in epilepsy, we have developed the IC-CoDE portal, an interactive web application that researchers can use to map the IC-CoDE taxonomy to their neuropsychological test scores. The portal enables rapid derivation of IC-CoDE cognitive phenotypes both at the single-patient level and at scale; researchers may upload a customized spreadsheet template populated with group data to obtain automatic phenotype assignments across large cohorts. In addition, the portal interface enables interactive pie-chart visualizations of phenotype distributions for own cohort exploration that can be dynamically stratified by user-specified demographic or clinical variables.

## Methods

### Development and Implementation of the IC-CoDE Portal

The IC-CoDE portal was developed as a fully browser-based web application using the Shiny framework in RStudio (R version 4.4.0; https://shiny.rstudio.com/). Shiny translates user inputs into HTML, performs server-side data processing in R, and renders results in the browser, enhanced with CSS themes and JavaScript widgets. The application is compatible with all major web browsers, including mobile devices. Portals are hosted by the Cleveland Clinic and deployed on Google Cloud services using self-contained Docker images for scalability and portability. The R Shiny code is packaged into a docker image, leveraging rocker/shiny:4.4.2 as the image base. This is then deployed to a Docker instance running on Linux. Visualizations are generated using the ggplot2 (https://ggplot2.tidyverse.org/) and plotly (https://plot.ly/r/) R libraries. The source code for the portal is openly available on the respective GitHub repository (https:github.com/TobiasBruenger/IC-Code-Portal), and a tutorial video demonstrating navigation and functionality is provided on the portal website (https://ic-code-portal.ccf.org/).

Once the initial portal build was complete, it was thoroughly reviewed and tested by all members of the study team, including seven clinical neuropsychologists specializing in epilepsy (R.M.B., K.A., J.K.J., S.J.S., A.R., C.R.M., B.P.H.). This included validity testing, comparing manual IC-CoDE phenotyping to portal-generated phenotyping. Once this process was complete, a link to the portal test site, along with an anonymous REDCap survey to rate aspects of the site, was sent to members of the ILAE Neuropsychology Task Force, the INS Epilepsy Special Interest Group, and researchers with previously published research on the IC-CoDE. Five neuropsychologists accepted the invitation to beta test the portal site. All reported that the layout and design were organized and user-friendly and the instructions on how to use the website were clear. One individual reported server connection issues and one had difficulty navigating the calculator itself. Four individuals provided information about what they liked best about the site (e.g., easy to use, organized cognitive domains, layout/design organized and user friendly, flexibility in terms of tests and types of scores) and four provided suggestions for improvement. Modifications and improvements were made to the portal to address the issues raised by the individuals who beta tested the site (i.e., clarity regarding test versions, providing a brief list of test instructions for ease of reference, labeling of graphs). It was then re-evaluated by the seven clinical neuropsychologists on the study team before the final IC-CoDE Portal website was launched at https://ic-code-portal.ccf.org/.

### Equipment Required

Use of the portal requires a computing device with an internet browser that has Javascript enabled. The following browsers have been tested: Google Chrome, Firefox, Safari, and Internet Explorer. In order to generate phenotypes using group data, spreadsheet software, such as Microsoft Excel, Libre Office Calc, Google Sheets, or Apple Numbers, is also required.

### Instructions for Portal Use

The IC-CoDE Web Portal can be accessed at https://ic-code-portal.ccf.org/. Users have the option of generating IC-CoDE cognitive phenotypes for an individual patient/participant or for a group of patients/participants using cognitive data obtained from neuropsychological evaluations. Detailed instructions along with an instructional video are provided under the “Instructions for Use” tab on the Web Portal and will only briefly be summarized here.

#### Calculating IC-CoDE Phenotype for an Individual Patient/Participant

There are four steps to calculating IC-CoDE phenotype(s) for a single patient/participant ***(Figure 1)***. *First*, users click on the “Individual” tab and select the threshold(s) they would like to use in generating IC-CoDE phenotype(s) (i.e., <-1.0, -1.5 and/or -2.0 standard deviations below the normative mean) and the scale to which they would like data entry to default (i.e., standard score, scaled score, T-score, z-score, percentile score). *Second*, users employ a series of drop-down menus, organized by cognitive domain, to enter cognitive tests and scores. Each cognitive domain contains several sub-categories (e.g., naming and fluency in the Language Domain; set-shifting, problem-solving, and response inhibition in the Executive Function Domain) to help guide the selection of measures that tap into different functions within each cognitive domain as recommended in the original IC-CoDE publication^10^. These sub-categories are provided only as a guide (i.e., they do not impact the phenotype classifications); users have the option of entering whatever tests they desire within each cognitive domain. However, it is recommended that users include tests of different types within a domain whenever possible (e.g., naming and fluency tasks rather than two fluency tasks in the Language Domain). There must be at least two tests within at least four cognitive domains for the calculator to generate a phenotype. Clicking the 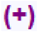 symbol next to the sub-category populates common cognitive tests for that domain and sub-category. The user can enter a standardized test score for a populated test or enter the name of a different cognitive test. Next the user indicates on what scale the score is measured (e.g., t-score). If the entered score is greater than 2 standard deviations above or below selected scale mean, a small warning sign 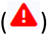 appears to the right of the test row. If the user hovers over the sign 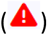, they are encouraged to review the score and scale to ensure that they are entered correctly. This is simply a prompt to check for errors; the calculator will still work if this warning button is present. Importantly, if the user enters data and then closes the sub-category (i.e., clicks the 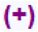 sign), the entered data will delete. So, users are encouraged to leave all tests lists expanded until they have generated the cognitive phenotype. *Third*, users may enter scores on screening measures of depression, anxiety, and/or behavior if they would like the IC-CoDE phenotype output to include this additional information. However, data entry under “MOOD AND BEHAVIOR” is optional and not required to generate a cognitive phenotype. Currently, only the populated measures can be used under the “MOOD AND BEHAVIOR” button. The user can hover over the 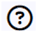 symbol to see how each measure is scored. *Finally*, after all test scores have been entered, the user clicks on the “SUBMIT ENTRIES” button to automatically generate the IC-CoDE cognitive phenotype(s). ***Figure 2*** shows a representative example of the IC-CoDE calculator output for an individual patient/participant. The user can download the results by selecting “DOWLOAD HTML”, modify entered data and re-run phenotypes by clicking “SUBMIT ENTRIES,” or select “RESET” to erase all entered data and start anew.

**Figure 1.**
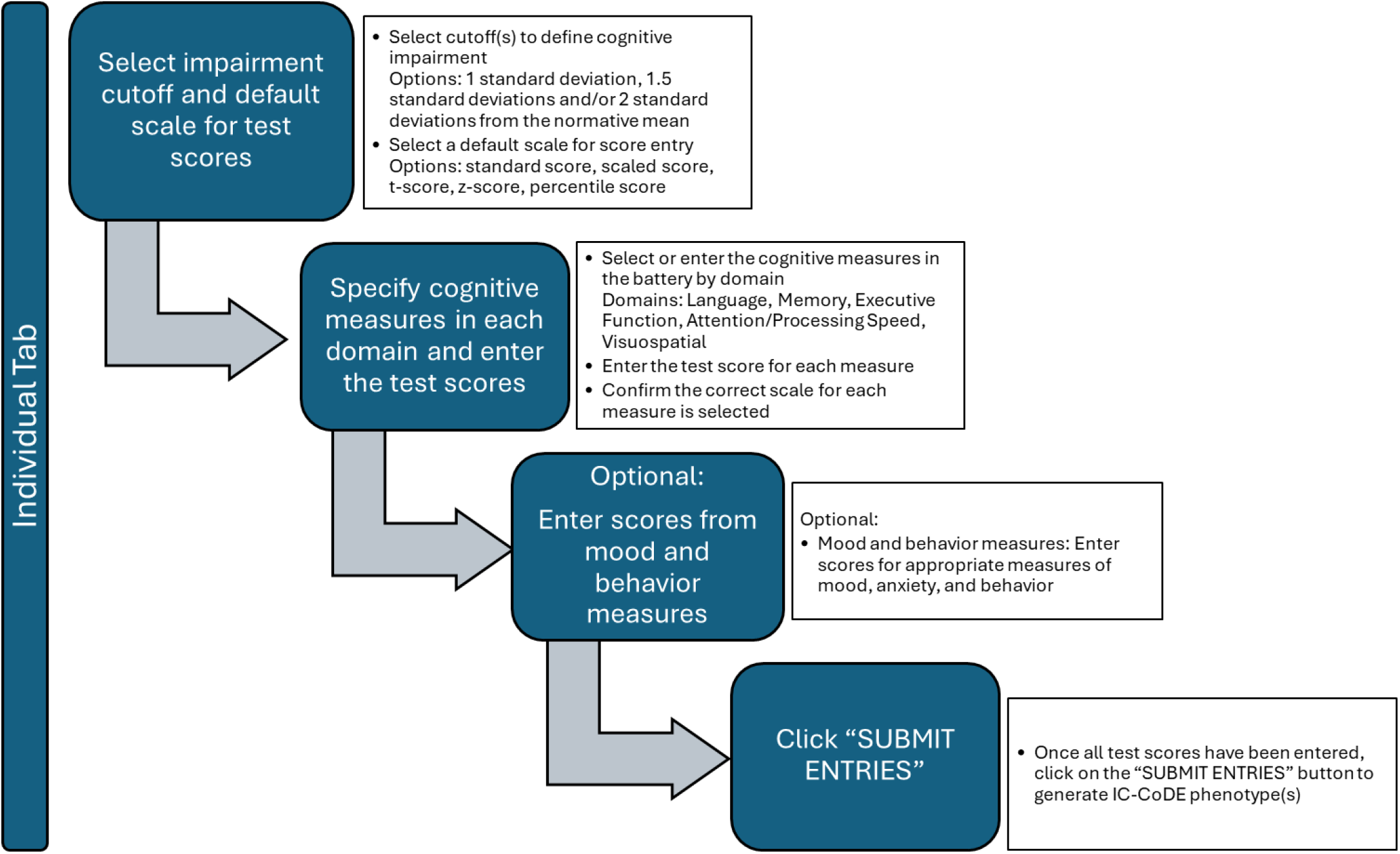
Four steps to generate IC-CoDE phenotype(s) for an individual patient/participant. The individual IC-CoDE calculator is accessed from the IC-CoDE Web Portal home page under the “Calculator” tab. The patient/participant must have at least 2 test scores in at least 4 cognitive domains to generate an IC-CoDE phenotype.

**Figure 2.**
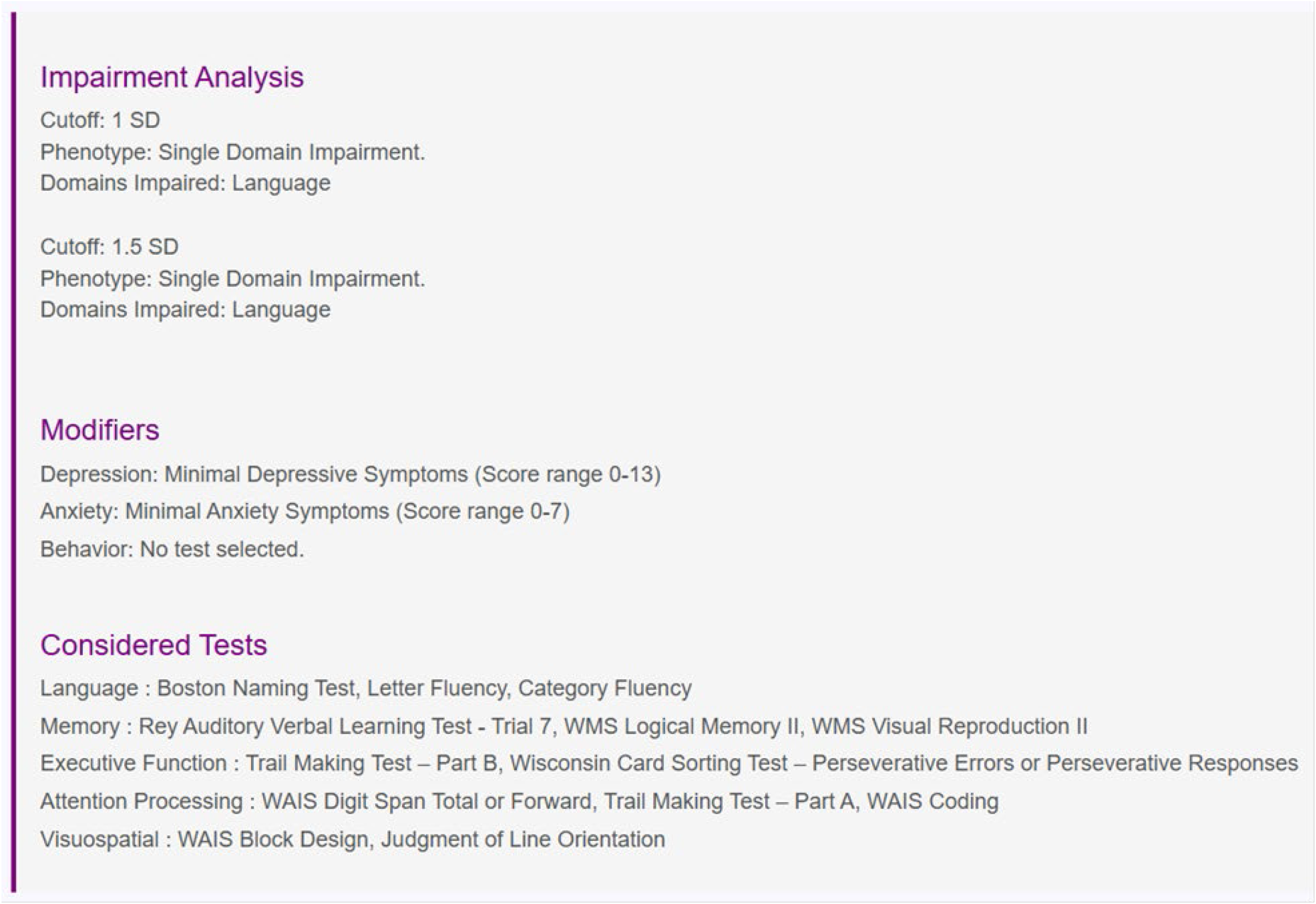
Example of IC-CoDE Calculator output for an individual patient/participant. The individual IC-CoDE calculator generates an on-screen output that includes an Impairment Analysis (i.e., the IC-CoDE phenotype and impaired domains for each cutoff selected), any Modifiers entered (i.e., depression, anxiety), and the Considered Tests within each domain used to generate the phenotype(s).

#### Calculating IC-CoDE Phenotype for a Group of Patients/Participants

There is a two-step process to calculate IC-Code phenotypes for a group of patients/participants in which the user creates a template and then uploads the data into the portal ***(Figures 3A*** *and* ***3B)***. To create the template, select the “Group” tab. *First*, as with the individual calculator described above, select the default scale for data entry. *Second*, users click on the “Create Template” tab and the same series of drop-down menus, organized by cognitive domain, appear. Users can open each sub-category by clicking the and select the cognitive tests in the neuropsychological battery by clicking the “Include” checkbox. If needed, you can change the type of standardized test score. As with the Individual calculator, the sub-categories are provided only as a guide (i.e., they do not impact the phenotype classifications); users have the option of selecting or entering whatever tests they desire within each cognitive domain. Although, as noted previously, it is recommended that users include tests of different types of abilities within a domain whenever possible (e.g., attention and processing speed tasks in the Attention/Processing Speed Domain rather than two measures of Attention). As with the individual calculator, there must be at least two tests within each of the cognitive domains and at least four cognitive domains for the calculator to generate a phenotype. *Third*, users may select optional mood and behavior measures and/or filters. As with the individual calculator, only the populated mood and behavior measures can currently be used. The scoring of the provided mood and behavior measures is available under the Individual calculator tab. With the group data, users may also create filter categories they may want to use when analyzing their phenotype data (e.g., demographic, disease, surgical variables). Several examples of common filters are provided, but the user has the option of entering any filter by simply typing over an existing filter label or the “Other (specify)” label. For example, in a research study an investigator may want to examine differences between patients with frontal and temporal lobe epilepsy, so they may choose to add “Site of seizures” as a filter, or they may want to examine the relationship between cognitive phenotypes and epilepsy variables and choose to add “Age at seizure onset” as a filter. Users must check the “Include” box to the right of each cognitive measure, mood or behavior measure, and filter they would like included in the database template. Measures entered without a check on the “Include” box will not be included in the template and in the subsequent phenotyping output. Before downloading, users should ensure that each test has the correct scale based on their data and adjust the scale if needed. *Fourth*, users click on the “DOWNLOAD GROUOP TABLE TEMPLATE” button. This downloads a file template in .xlsx format. The first row of the template contains the selected tests and filters as variable names in a special format for the IC-CoDE Calculator. The first column supplies the user with a legend for text color. When entering **de-identified group data** into the template, it is imperative that the first column and variable names in the first row not be altered from how they were downloaded. If there is any modification to the variable names, the IC-CoDE Calculator will not correctly generate cognitive phenotypes. It is also important to note that values entered into the template that may be out of the expected range (2 standard deviations above or below the mean) will turn red to signal to the user that they should double check score entry and score type (i.e., scaled, standard) to ensure the accuracy of the entered data. This is simply a prompt to check for errors; the calculator will still work if red scores remain present when the template is uploaded back into the IC-CoDE Portal. Only numbers can be entered under the cognitive test and mood or behavior score columns. Text can be entered under the filters. While no information entered or uploaded into the portal is stored, the user should not enter or upload any protected health information (e.g., names, dates of birth, addresses, medical record numbers) into the data template. Column B of the template allows the user to enter an optional StudyID.

**Figure 3.**
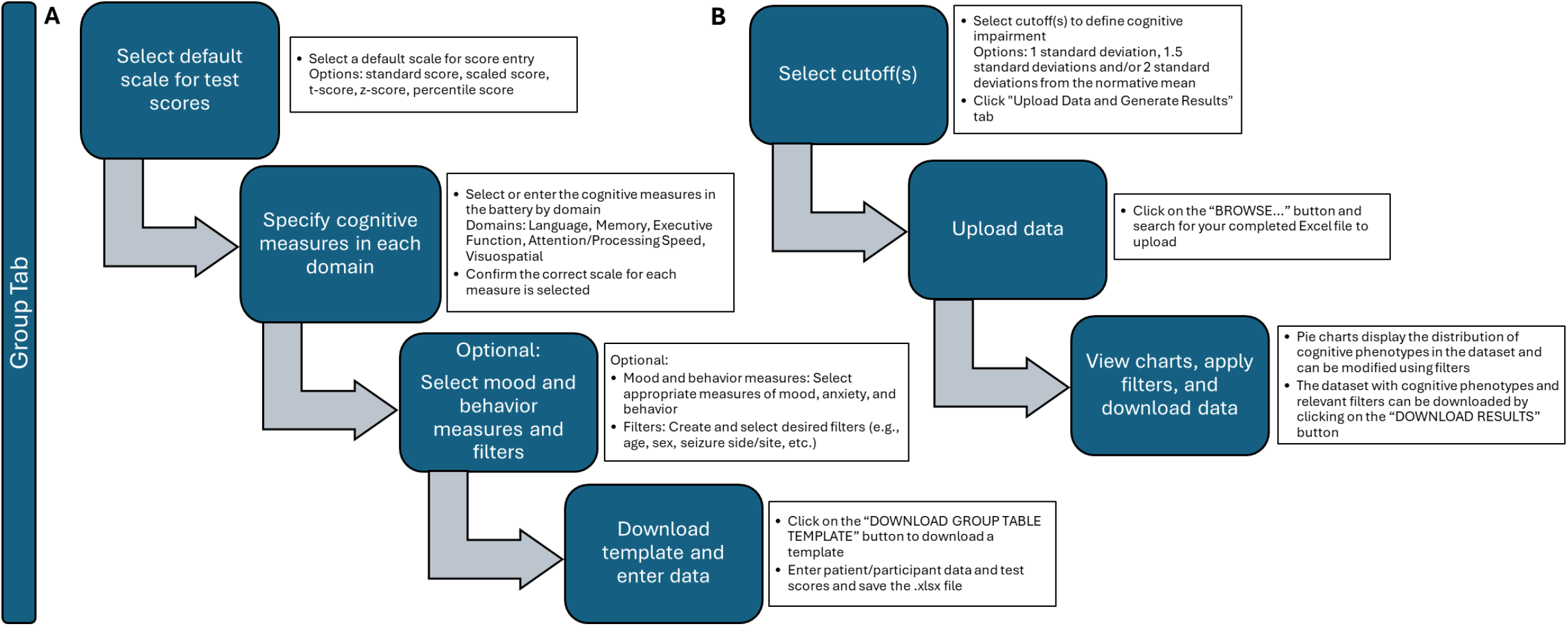
Generating IC-CoDE phenotypes for a group of patients/participants. **A)** The four steps to prepare a data template to enter group data. **B)** The three steps to upload completed dataset and generate IC-CoDE phenotypes for a group of patients/participants. The IC-CoDE calculator is accessed from the IC-CoDE Web Portal home page under the “Calculator” tab. Each patient/participant must have at least 2 test scores in at least 4 cognitive domains to generate an IC-CoDE phenotype.

Once data entry into the template has been completed and saved as an Excel file (i.e., .xlxs), the user can return to the IC-CoDE Portal and go directly to the “Group” Tab and “Upload Data and Generate Results” Tab. The user then selects the cutoff(s) for IC-CoDE Phenotypes. Published studies to date have typically used 1.0 or 1.5 standard deviation cutoffs, with the 1.5 cutoff used most frequently^10,12,15,17,18^.

Next, the user clicks on the “BROWSE” button at the bottom of the page and selects the completed Excel file for upload. Once the file has been uploaded, pie chart(s) demonstrating the distribution of IC-CoDE phenotypes in the dataset will be displayed on the portal site (***Figure 4***). The generated pie chart(s) are interactive. The user can modify the cut off(s) of the presented phenotypes at any time. Hovering over the pie slices displays information about the number of individuals in the cognitive domain and supplies details on the base rate of each type of domain impairment (e.g., single domain: language, single domain: memory, bi-domain: language and memory, bi-domain: memory and executive function). The display results can also be altered by applying filters (e.g., sex, seizure side). Just below the pie charts, there is an option to “DOWNLOAD RESULTS.” Clicking this button will initiate download of a dataset in a .csv file containing the IC-CoDE phenotypes for each patient/participant and the domains that were impaired along with any relevant filters that were included in the uploaded data file. This dataset can then be used to facilitate offline analysis of cognitive data by users. Data entered or uploaded into the portal is not saved, but it is best practice to avoid entering any identifying information. Because the portal does not store data, it is recommended to download the results immediately before the session times out.

**Figure 4.**
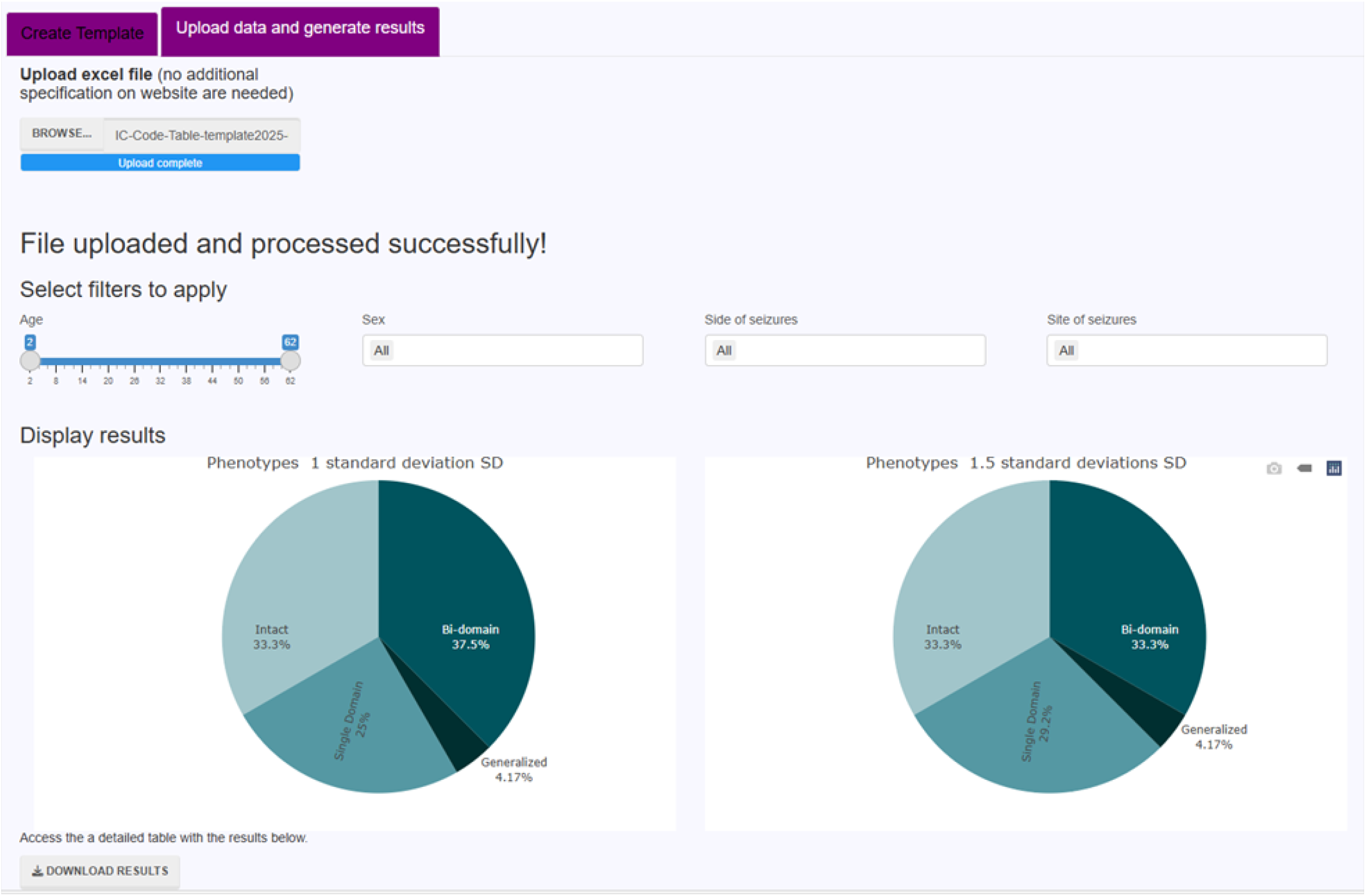
Pie Charts Demonstrating Distribution of IC-CoDE Phenotypes

## Conclusions

The IC-CoDE portal operationalizes a consensus taxonomy for cognitive phenotyping in epilepsy, transforming what was a labor-intensive, error-prone procedure into a rapid, reproducible workflow accessible to investigators worldwide. By automating phenotype assignment across heterogeneous neuropsychological batteries, the web application markedly lowers the barrier to multicenter data harmonization, thereby facilitating well-powered studies capable of dissecting the neural substrates and clinical sequelae of cognitive dysfunction in epilepsy.

Beyond phenotype generation, the portal’s built-in visual analytics and download options promote transparent data exploration and seamless integration with downstream statistical pipelines. Early user testing demonstrated high usability and highlighted iterative improvements, underscoring the value of community-driven development. Further, anecdotally, neuropsychology members of the development team have noted that use of the portal is phenomenal in terms of the accuracy and speed with which they can generate phenotypes compared to the manual methods they used in research prior to portal development, particularly in large datasets with diverse cognitive batteries. Its ability to calculate phenotypes using multiple cut scores simultaneously, all while factoring in individual differences in the tests administered, make it an ideal tool for cognitive phenotyping. The IC-CoDE portal can also accelerate ongoing multi-center and international research efforts (e.g., ENIGMA-Epilepsy) where imaging and genetic data have already been harmonized, but neuropsychological data have not. Future, multi-center efforts are already underway to determine the optimal procedures for phenotype generation (e.g., using all available tests vs. limiting to two tests per domain for phenotype generation; using ipsative vs. normative approaches to define cognitive impairment; inclusion of a single memory domain or separate domains for verbal and visuospatial memory). While IC-CoDE continues to be for research only, it is hoped that these studies and others may eventually lead to an optimal cognitive phenotype model that can be translated to the clinical context. The IC-CoDE portal has a Publications tab where up-to-date references are provided as research advances. There is also a FAQ tab, which we plan to update if guidelines are adjusted based on future research.

Looking forward, planned IC-CoDE portal enhancements include integration of an Application Programming Interface (API) for programmatic batch processing and linkage with imaging and genetic repositories to support multimodal phenotype–biomarker analyses. Collectively, the IC-CoDE portal represents a scalable, open-access resource poised to accelerate discovery in epilepsy neuropsychology and to inform precision-medicine initiatives targeting cognition-related comorbidities.

## Data Availability

Data sharing is not applicable as no datasets were generated or analyzed during the current study.

## Acknowledgments

The authors thank the members of the ILAE Neuropsychology Task Force as well as those individuals in the epilepsy neuropsychology research community who have collaborated with us on IC-CoDE-related projects and the development of the IC-CoDE portal. We would also like to thank the members of the epilepsy neuropsychology community who tested the beta version of the IC-CoDE portal and provided us with valuable feedback. Finally, we are grateful for the assistance of Mark St. John and Brook Hurd, who were instrumental in facilitating launch of the portal.

## Author Contributions

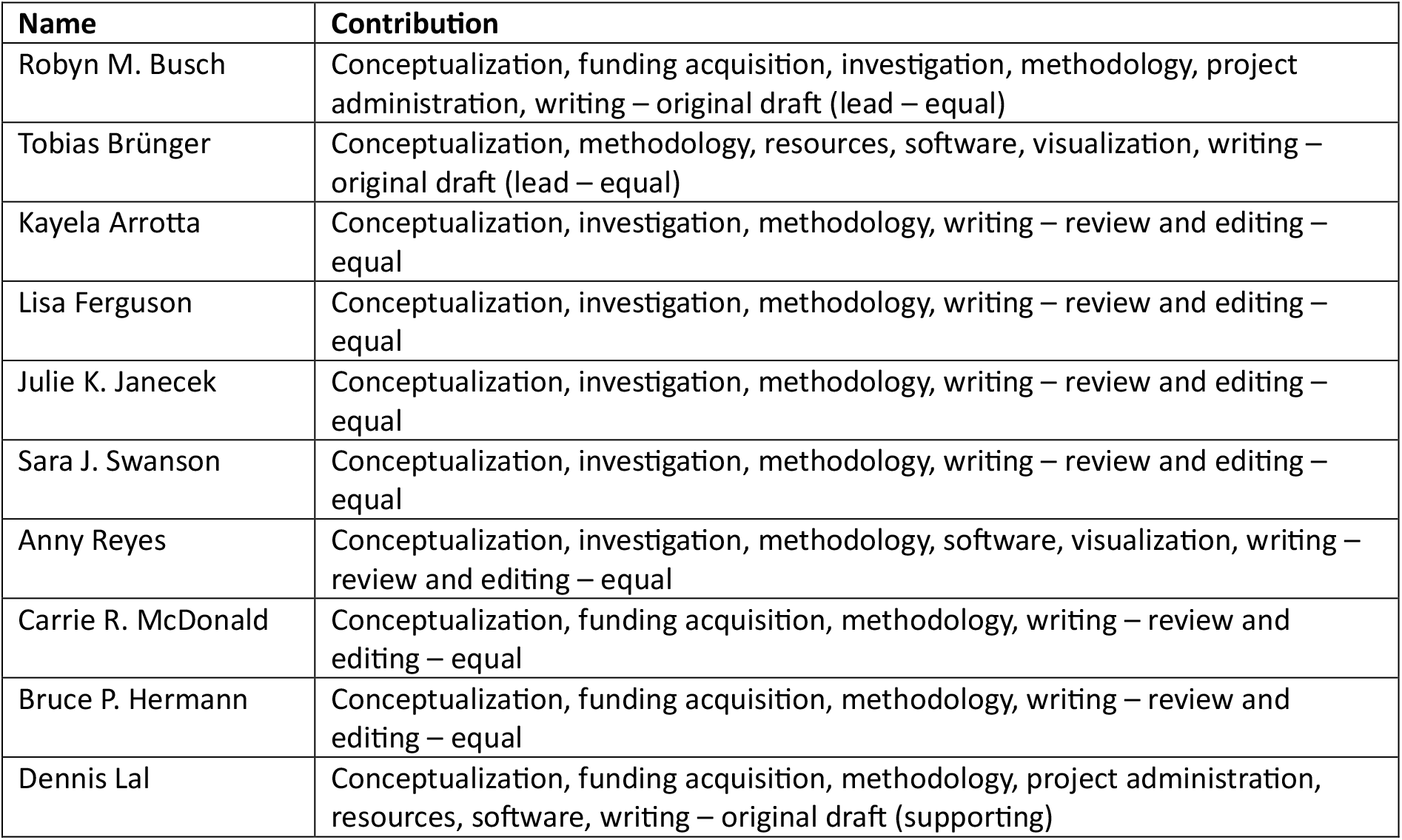

## Ethical Publication Statement

The authors confirm that we have read the Journal’s position on issues involved in ethical publication and affirm that this report is consistent with those guidelines.

## References

1. Giovagnoli AR, Parente A, Tarallo A, Casazza M, Franceschetti S, Avanzini G. Self-rated and assessed cognitive functions in epilepsy: impact on quality of life. Epilepsy Res. 2014; 108(8):1461–8.

2. McAuley JW, Elliott JO, Patankar S, Hart S, Long L, Moore JL, et al. Comparing patients’ and practitioners’ views on epilepsy concerns: a call to address memory concerns. Epilepsy Behav EB. 2010; 19(4):580–3.

3. Loring DW. History of neuropsychology through epilepsy eyes. Arch Clin Neuropsychol Off J Natl Acad Neuropsychol. 2010; 25(4):259–73.

4. Norman M, Wilson SJ, Baxendale S, Barr W, Block C, Busch RM, et al. Addressing neuropsychological diagnostics in adults with epilepsy: Introducing the International Classification of Cognitive Disorders in Epilepsy: The IC CODE Initiative. Epilepsia Open. 2021; 6(2):266–75.

5. Hermann B, Conant LL, Cook CJ, Hwang G, Garcia-Ramos C, Dabbs K, et al. Network, clinical and sociodemographic features of cognitive phenotypes in temporal lobe epilepsy. NeuroImage Clin. 2020; 27:102341.

6. Dabbs K, Jones J, Seidenberg M, Hermann B. Neuroanatomical correlates of cognitive phenotypes in temporal lobe epilepsy. Epilepsy Behav EB. 2009; 15(4):445–51.

7. Reyes A, Kaestner E, Bahrami N, Balachandra A, Hegde M, Paul BM, et al. Cognitive phenotypes in temporal lobe epilepsy are associated with distinct patterns of white matter network abnormalities. Neurology. 2019; 92(17):e1957–68.

8. Kaestner E, Reyes A, Macari AC, Chang Y-H, Paul BM, Hermann BP, et al. Identifying the neural basis of a language-impaired phenotype of temporal lobe epilepsy. Epilepsia. 2019; 60(8):1627–38.

9. Struck AF, Boly M, Hwang G, Nair V, Mathis J, Nencka A, et al. Regional and global resting-state functional MR connectivity in temporal lobe epilepsy: Results from the Epilepsy Connectome Project. Epilepsy Behav EB. 2021; 117:107841.

10. McDonald CR, Busch RM, Reyes A, Arrotta K, Barr W, Block C, et al. Development and application of the International Classification of Cognitive Disorders in Epilepsy (IC-CoDE): Initial results from a multi-center study of adults with temporal lobe epilepsy. Neuropsychology. 2022; .

11. Shah U, Rajeshree S, Sahu A, Kalika M, Ravat S, Reyes A, et al. Cross-cultural application of the International Classification of Cognitive Disorders in Epilepsy (IC-CoDE): Cognitive phenotypes in people with temporal lobe epilepsy in India. Epilepsia. 2024; 65(8):2386–96.

12. Arrotta K, Swanson SJ, Janecek JK, Hamberger MJ, Barr WB, Baxendale S, et al. Application of the International Classification of Cognitive Disorders in Epilepsy (IC-CoDE) to frontal lobe epilepsy using multicenter data. Epilepsy Behav EB. 2023; 148:109471.

13. Bingaman N, Ferguson L, Thompson N, Reyes A, McDonald CR, Hermann BP, et al. The relationship between mood and anxiety and cognitive phenotypes in adults with pharmacoresistant temporal lobe epilepsy. Epilepsia. 2023; 64(12):3331–41.

14. Busch RM, Dalton JE, Jehi L, Ferguson L, Krieger NI, Struck AF, et al. Association of Neighborhood Deprivation with Cognitive and Mood Outcomes in Adults with Pharmacoresistant Temporal Lobe Epilepsy. Neurology. 2023; 100(23):e2350–9.

15. Reyes A, Salinas L, Hermann BP, Baxendale S, Busch RM, Barr WB, et al. Establishing the cross-cultural applicability of a harmonized approach to cognitive diagnostics in epilepsy: Initial results of the International Classification of Cognitive Disorders in Epilepsy in a Spanish-speaking sample. Epilepsia. 2023; 64(3):728–41.

16. Miron G, Müller PM, Hohmann L, Oltmanns F, Holtkamp M, Meisel C, et al. Cortical Thickness Patterns of Cognitive Impairment Phenotypes in Drug-Resistant Temporal Lobe Epilepsy. Ann Neurol. 2024; 95(5):984–97.

17. Almane DN, Busch RM, Ferguson L, Arenivas A, Reyes A, McDonald CR, et al. Application of the international classification of cognitive disorders in epilepsy (IC-CoDE) to youths with new and recent onset epilepsies. Epilepsy Behav EB. 2025; 171:110606.

18. Ferguson L, Arenivas A, Arrotta K, Almane D, Jones J, Reyes A, et al. Application of the International Classification of Cognitive Disorders in Epilepsy (IC-CoDE) to youths with drug-resistant epilepsy. Epilepsy Behav EB. 2025; 171:110607.

19. Reyes A, Hermann BP, Prabhakaran D, Ferguson L, Almane DN, Shih JJ, et al. Validity of the MoCA as a cognitive screening tool in epilepsy: Are there implications for global care and research? Epilepsia Open. 2024; 9(4):1526–37.

20. Sarkis RA. Update in Progress: Cognitive Phenotypes in Temporal Lobe Epilepsy. Epilepsy Curr. 2023; 23(6):363–5.

21. Widdess-Walsh P. Breaking the CoDE of Cognitive Disorders in Epilepsy. Epilepsy Curr. 2023; 23(3):156–8.

